# The reproductive number *R*_0_ of COVID-19 based on estimate of a statistical time delay dynamical system

**DOI:** 10.1101/2020.02.17.20023747

**Authors:** Nian Shao, Jin Cheng, Wenbin Chen

## Abstract

In this paper, we estimate the reproductive number *R*_0_ of COVID-19 based on Wallinga and Lipsitch framework [11] and a novel statistical time delay dynamic system. We use the observed data reported in CCDC’s paper to estimate distribution of the generation interval of the infection and apply the simulation results from the time delay dynamic system as well as released data from CCDC to fit the growth rate. The conclusion is: Based our Fudan-CCDC model, the growth rate *r* of COVID-19 is almost in [0.30, 0.32] which is larger than the growth rate 0.1 estimated by CCDC [9], and the reproductive number *R*_0_ of COVID-19 is estimated by 3.25 ≤ *R*_0_ ≤ 3.4 if we simply use *R* = 1 + *r ∗ T*_*c*_ with *T*_*c*_ = 7.5, which is bigger than that of SARS. Some evolutions and predictions are listed.

## 1 One formula for the distribution of generation interval

The serial interval distribution presented in the paper “Early transmission dynamics in Wuhan, China, of novel coronavirus-infected peneumonia” [9] by the researches in Chinese Center for Disease Control and Prevention(CCDC) et al, which is defined as the duration between symptom onset insuccessive cases in the transmission chain in Fig.2B of [9]. We assume that the distribution is Gamma distribution, i.e., the corresponding probability density function is

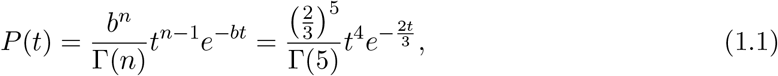

where the parameters are *n* = 5 and 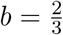. So the mean of the distribution is 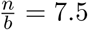 which is exactly equal to the number announced in [9], where they claimed that the mean serial interval of COVID-19 is 7.5 days (95% CI, 5.3 to 19). Besides the mean, comparison of (1.1) with Fig2B is more convince. The formula is so beautiful, that **we believe it must be true**. The other distributions are also given in our recent report [10].

**Figure 1:**
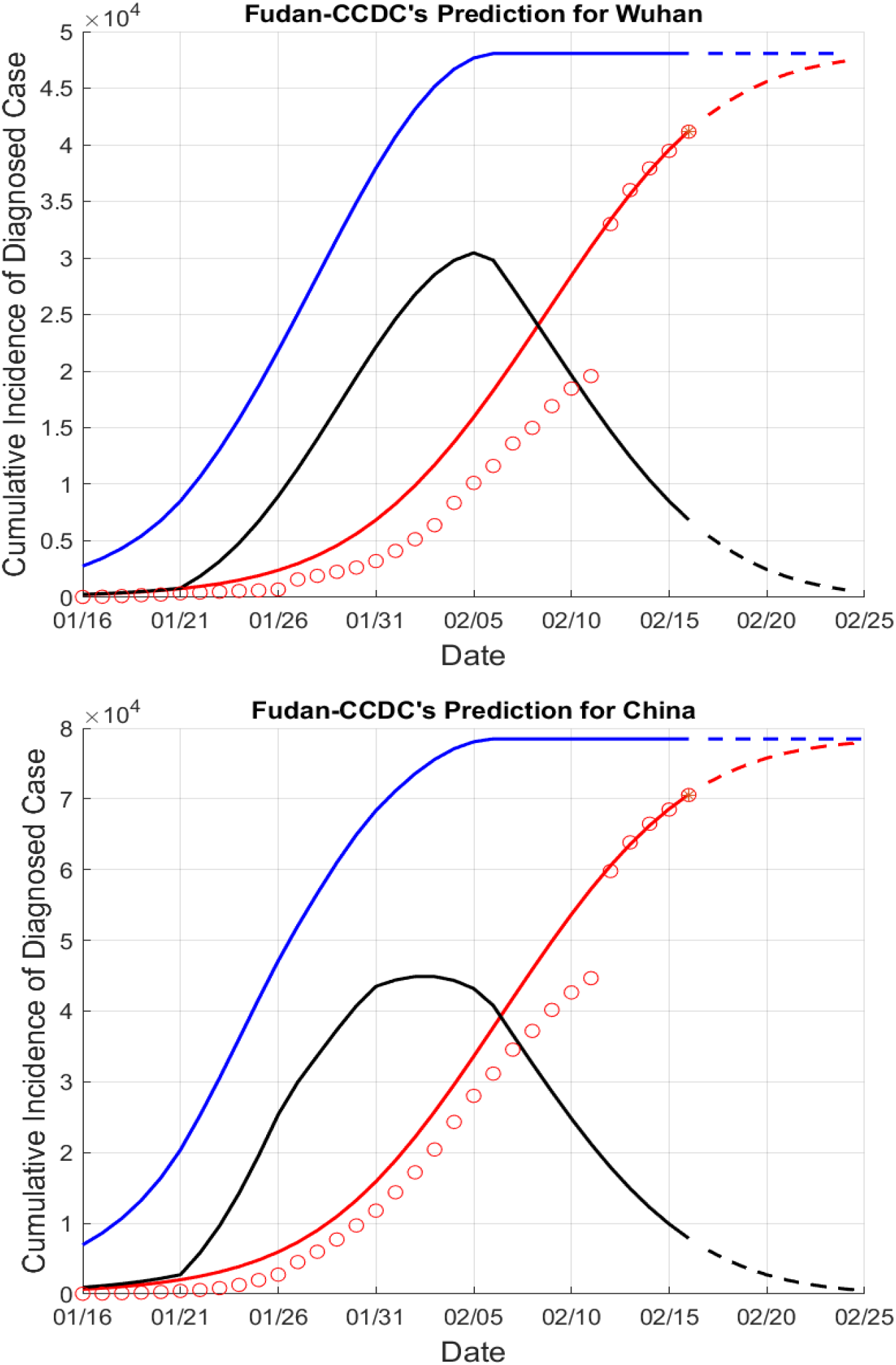
Evolution of COVID-19

**Figure 2:**
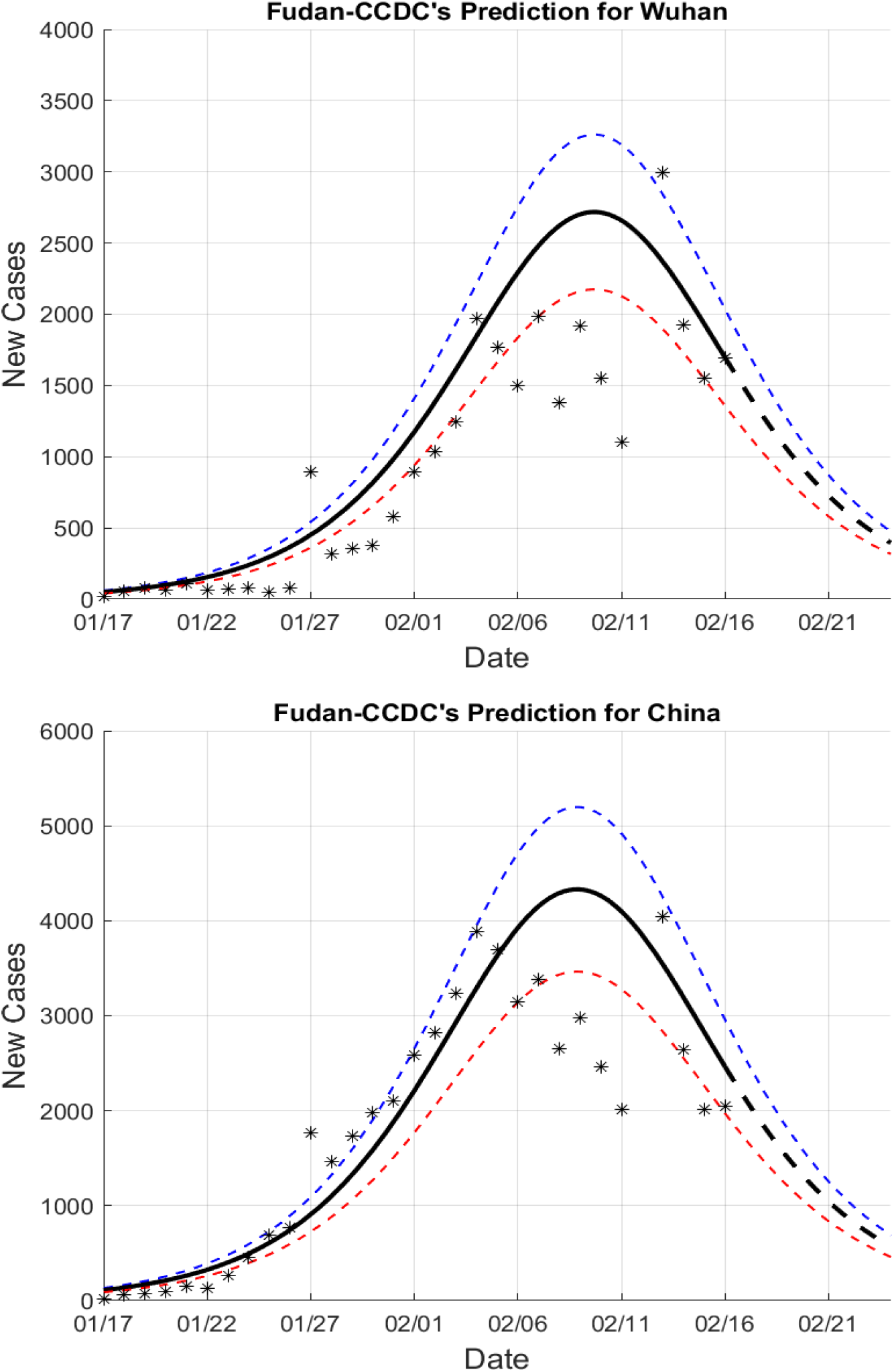
New cases

**Figure 3:**
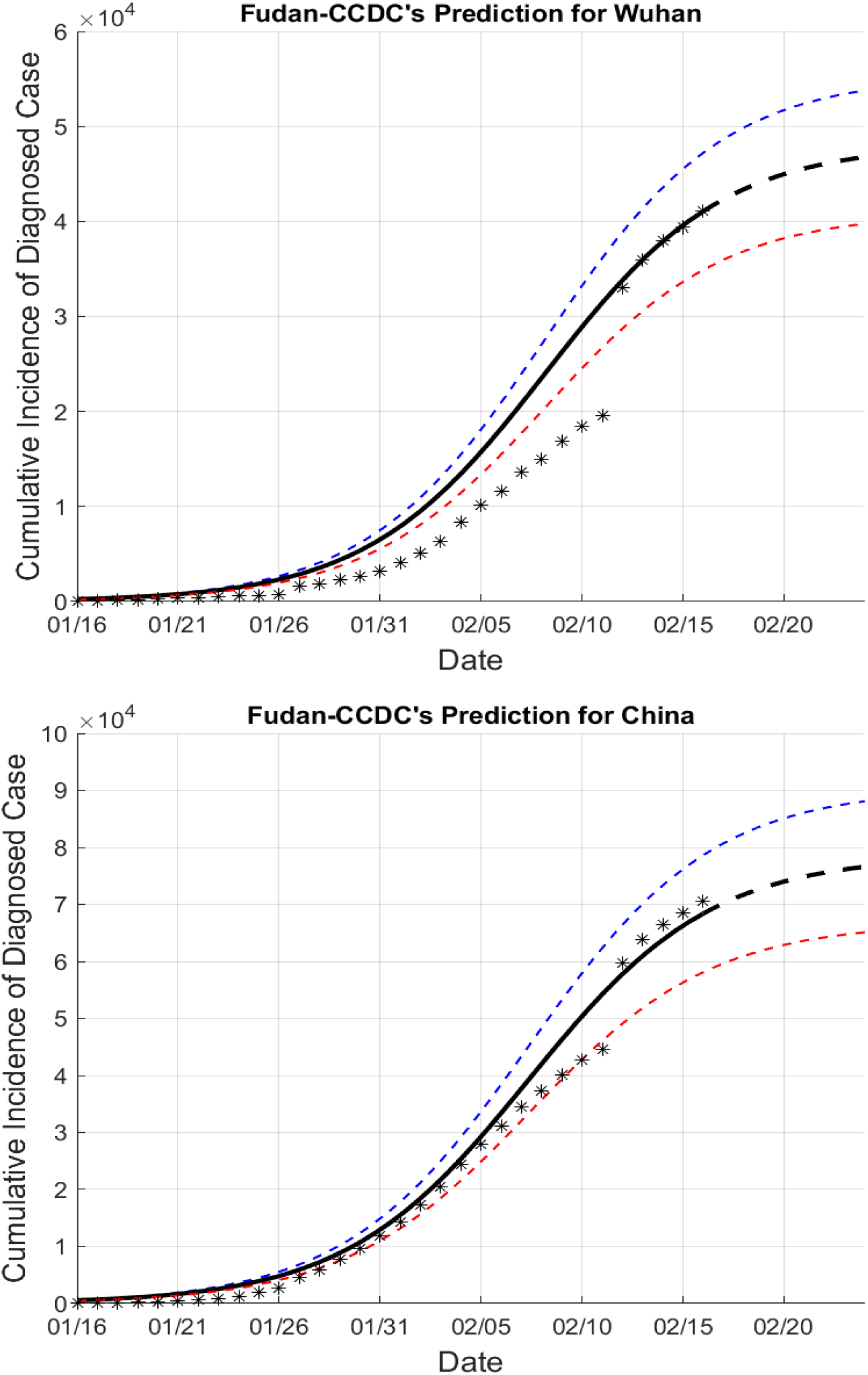
Cumulative incidence of diagnosed case

We think this distribution just as the one of generation interval mentioned in [11]. Now we can determine the reproductive number *R*_0_ of COVID-19 if we know the growth rate *r* of COVID-19 by

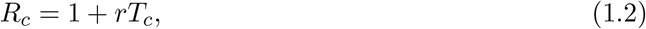

where *T*_*c*_ = 7.5 can be obtained from (1.1). Before we discuss how to get the estimate of the growth rate *r* of COVID-19, let us review the elegant framework by Wallinga and Lipsitch [11].

## 2 Wallinga and Lipsitch’s framework

According to the framework of Wallinga and Lipsitch [11], in order to obtain the reproductive number *R*_0_, some variables and distributions should be known based on the data before we use their uniform framework:

- the growth rate of the epidemic *r*;
- the mean generation interval of the infection *T*_*c*_;
- or the generation interval distribution of the infection *g*(*t*).

We just mention that the mean generation interval *T*_*c*_ is the first moment of the generation interval distribution *g*(*t*):

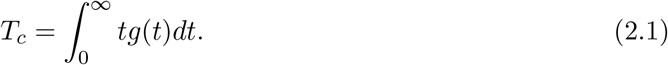

If *r* and *T*_*c*_ are computed, the value of the reproductive number *R*_0_ can be simply estimated through a linear equation([11, 1, 8, 4, 5]):

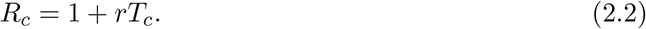

And if *r* and *g*(*t*) are estimated, *R*_0_ can be computed as

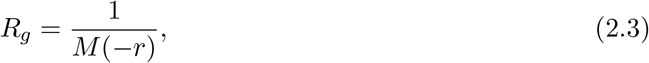

where *M* (*z*) is the moment generating function of the distribution *g*(*t*):

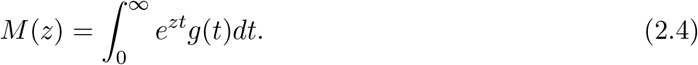

Wallinga and Lipstch’s processes are simple but powerful, which clearly demonstrate how generation intervals shape the relationship between growth rate and reproductive number. Specially, if *g*(*t*) is Gamma distribution with parameters (*b, n*) as in [11], then we can have two estimates of *R*_0_, one is by (2.2)

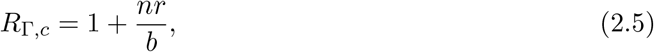

and the another is by (2.3)

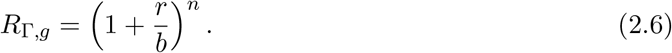

Based on the formula of the generation interval, we know that *b* = ^2^ and *n* = 5, *R*_Γ,*g*_ will be too sensitive on *r*, and we will choose (1.2) to obtain *R*_0_, and our main focus is to get an accurate and stable estimate for the growth rate *r*. The modified second order approximation to (2.6) is also important and has been widely used[11, 6].

In [11], Wallinga and Lipsitch point out that if the epidemic model is assumed to be susceptible-exposed-infectious-recovered(SEIR) model, then the generation interval distribution is Gamma distribution.

So we can obtain our first conclusion.

### Conjecture 2.1

*The epidemic model for COVID-19 is SEIR if CCDC’s data are correct*.

The above conclusion may be interpreted by that CCDC assumed that the epidemic model should be SEIR.

Now we introduce how we can estimate the growth rate *r*.

## 3 Fudan-CCDC model

Our group has developed some models for the growth rate of COVID-19 (TDD-NCP models [2, 13] and Fudan-CCDC models[10]) after Jin suggested to use the time delay model to fit the real data [2]. Because of the outstanding agreement with data from CCDC and the estimations on the growth rate are very stable, we decide to use the following Fudan-CCDC model. Other models also work very well to fit the CCDC data.

Firstly, Let us introduce some variables:

- *I*(*t*): (the distribution of) the cumulative number of cases in the infected stage at time *t*;
- *J* (*t*): (the distribution of) the cumulative ones in hospitalization stage at time *t*;
- *G*(*t*): (the distribution of) the number of infected and isolated but yet diagnosed cases at time *t*. They are infected in fact, but are not confirmed by the hospital, then not appeared in the infected list of CCDC;
- *I*_0_(*t*): *I*_0_(*t*) = *I*(*t*) *− J* (*t*) *− G*(*t*).

We further make some assumptions on the transition probability functions *f*_2_(*t*), *f*_3_(*t*) and *f*_4_(*t*):

- *f*_2_(*t*): the transition probability from infection to illness onset;
- *f*_3_(*t*): the transition probability from illness onset to hospitalization;
- *f*_4_(*t*): the transition probability from infection to hospitalization, which can be calculated via the convolution of *f*_2_(*t*) and *f*_3_(*t*).

We assume the log-normal distribution for *f*_2_(*t*) and the Weibull distrubution for *f*_3_(*t*)[10], and the distribution parameters can be estimated from CCDC by fitting the figures in [9]. In particular, two important parameters are used:

- *β*: denotes the growth rate, which is exactly the same as *r*: *β* = *r*.
- *ℓ*: denotes the quarantine strategy, for example, specially, we consider this parameter as the isolation rate. Large value of *ℓ* suggests that the government more likes to control the spreading of COVID-19.

Now we introduce our Fudan-CCDC model: *I*(*t*), *J* (*t*) and *G*(*t*) can be approximated by the following (statistical) dynamical system:

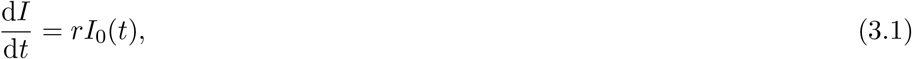

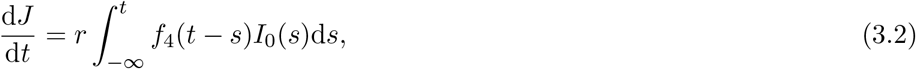

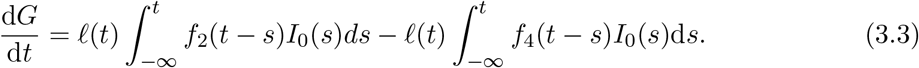

One can also use the discrete system with each step representing one day just as we have implemented in the code:

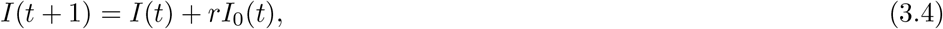

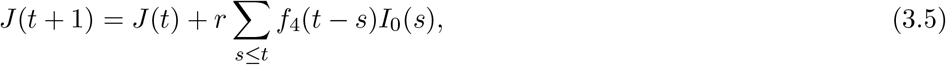

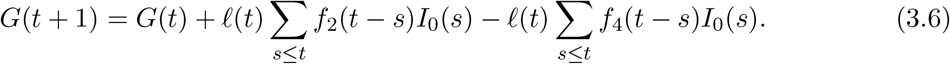

We will fit and estimate appropriate values of *β*(or *r*) and *ℓ* based on the cumulative diagnosed data from CCDC. Other versions of our Fudan-CCDC model are developing, specially, we are considering the stochastic version: that is to take random inputs into account, which is important to account for the influences from other regions. In math, we can easily generalize the model by random terms such as Brownian motion in the system, meanwhile the parameters have to be obtained from the real data. The another important version is to consider the interactions of two or more regions, Jin’s group have developed some models based on TTD-NCP model[3, 7]. These models will also be developed in need of disease control and prevention.

The simulation is implemented via Matlab with some additional constraints imposed. Here the results are summarized in Table 1 with the real data employed up to Feb 16, 2020.

**Table 1:**
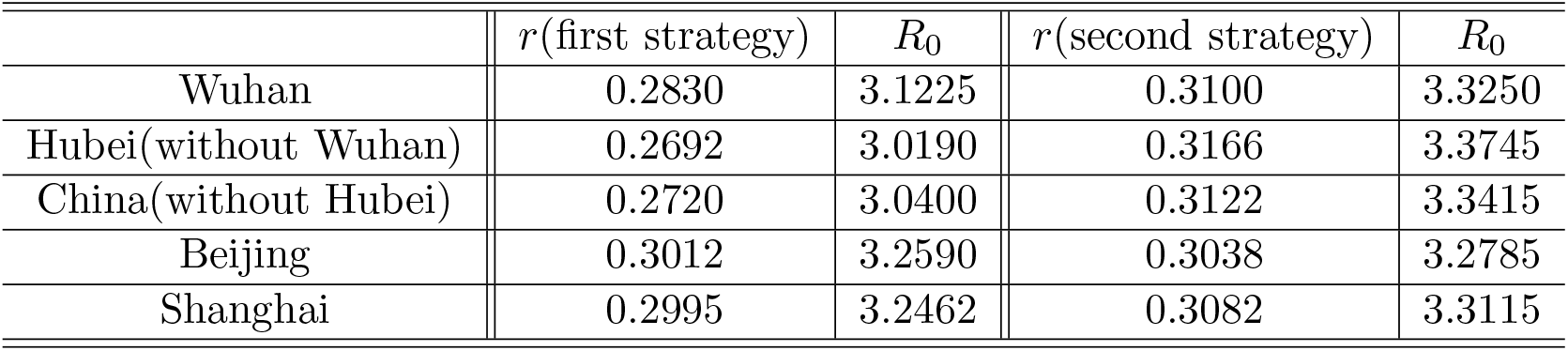
The growth rate *r* and the reproductive number *R*_0_

| | $r$(first strategy) | $R_0$ | $r$(second strategy) | $R_0$ |
| --- | --- | --- | --- | --- |
| Wuhan | 0.2830 | 3.1225 | 0.3100 | 3.3250 |
| Hubei(without Wuhan) | 0.2692 | 3.0190 | 0.3166 | 3.3745 |
| China(without Hubei) | 0.2720 | 3.0400 | 0.3122 | 3.3415 |
| Beijing | 0.3012 | 3.2590 | 0.3038 | 3.2785 |
| Shanghai | 0.2995 | 3.2462 | 0.3082 | 3.3115 |

In the Table 1, we use two kind of quarantine strategies introduced below. We are surprised that our code can get uniform estimators on the growth rate *r* and the reproductive number *R*_0_. **Two quarantine strategy:** We can observe that *ℓ* is distinct for different time stages at the different regions. Now we assume two quarantine strategies, one uses the following assumption:

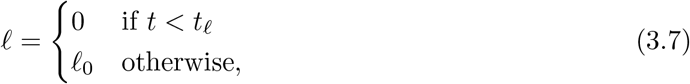

and one can use another strategy

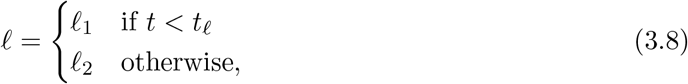

which means that the quarantine strategy was observably changed at the day *t_ℓ_*. We report the results in Table 2.

**Table 2:**
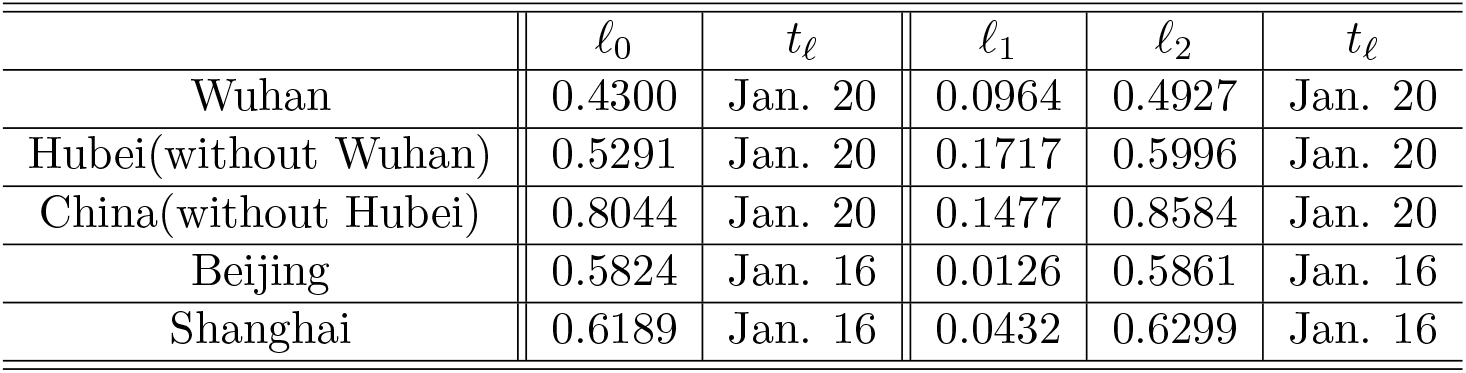
The isolation rate *ℓ* and the changed point *t_ℓ_*

**Table 3:** The initial infections *I*(*t*_0_) and the estimated initial date *t*_0_

| | parameter $I(t_0)$ | $t_0$(first strategy) | $t_0$(second strategy) |
| --- | --- | --- | --- |
| Wuhan | 4 | Dec. 19 | Dec. 19 |
| Hubei(without Wuhan) | 4 | Dec. 19 | Dec. 19 |
| China(without Hubei) | 4 | Dec. 19 | Dec. 19 |
| Beijing | 1 | Dec. 30 | Dec. 28 |
| Shanghai | 1 | Dec. 30 | Dec. 28 |

We also find that *ℓ* for the Diamond Princes Cruise Ship is *β* = 0.3182, *ℓ*_1_ = 0, *ℓ*_2_ = 0.3239. Moreover, the results from the model imply the case in the Diamond Princes Cruise Ship may be very DANGEROUS, since *ℓ* is below one key THRESHOLD, where the number of the infected persons will growth quickly. We also very worry about the situations in Japan. We WARN here that every CDC in every country should track the number of the infected persons, and immediately increase *ℓ* to one suitable level.

### Starting date *t*_0_

Our model can also track the initial date of the event *t*_0_ if *I*(*t*_0_) is fixed. It is REALLY important to find the initial starting date statistically correct in different regions.

## 4 Evolutions and predictions

Now we present some figures of the evolutions and predictions of the event computed from the model, The data used in the simulation is ONLY number of the cumulative diagnosed people of different regions from CCDC until Feb. 16, everyone can download it from many websites or APP, we find the data from WIND(like Bloomberg). And the second quarantine strategy is imposed.

### Evolutions

Here is the evolution of the cumulative number of the infected cases, cumulative number of cases in hospitalization *J* (*t*), and the cumulative number of infected and isolated but yet diagnosed cases *G*(*t*). According to our model, total number of infected cases will be **48060**(Wuhan), and **78480**(China). Based on this data, it is easy to estimate various data, specially the fatality rate can be estimated directly. We would like to point out that the rate calculated here will be varying once the quarantine strategy is changed as the rate depend on *ℓ*.

We also can obtain the cumulative incidence of diagnosed case and new cases that occurred in a given period which is displayed in Figure 2.

Some figures and other results have been submitted to Chinese Center for Disease Control and Prevention(CCDC), Wuhan.

## 5 Conclusions on the reproductive number

Based our Fudan-CCDC model, the growth rate *r* of COVID-19 is almost in [0.30, 0.32] which is larger than the growth rate 0.1 estimated by CCDC [9], and the reproductive number *R*_0_ of COVID-19 is estimated by 3.25 ≤ *R*_0_ ≤ 3.4 if we simply use *R*_0_ = 1 + *r ∗ T*_*c*_ with *T*_*c*_ = 7.5.

We have not compared the results with [12] and [14], since we are not very familiar with their methods.

## Data Availability

the data is from the annocements of Chinese Center for Disease Control and Prevention, and published papers.

## Ethics approval and consent to participate

The ethical approval or individual consent was not applicable.

## Availability of data and materials

All data and materials used in this work were publicly available.

## Consent for publication

Not applicable.

## Disclaimer

The funding agencies had no role in the design and conduct of the study; collection, management, analysis, and interpretation of the data; preparation, review, or approval of the manuscript; or decision to submit the manuscript for publication. Authors contributions All authors conceived the study, carried out the analysis, discussed the results, drafted the first manuscript, critically read and revised the manuscript, and gave final approval for publication.

## Authors contributions

The simulations are main implemented by Shao Nian and designed by Wenbin Chen. All authors conceived the study, carried out the analysis, discussed the results, drafted the first manuscript, critically read and revised the manuscript, and gave final approval for publication.

## Acknowledgements

We should truly thank Prof. Daqian Li and Prof. Guanhong Ding for their leading work on the SARS models, we are also sincerely grateful to every member in our group, special to Dr. Yu Jiang, Dr. Yue Ye, Dr. Boxi Xu, Dr. Yu Chen, Dr. Keji Liu, Miss Xinyue Luo, Dr. Ming Zhong, Dr. Xiang Xu and Prof. Shuai Lu and other group members. We also thank Yongzhen Wang and Prof. Shanjian Tang(Fudan), Gang Liang(China CDC in Wuhai, Hubei), our old friend Dr. Zhihua Shen(Fusion Fin Trade, fusionfintrade.com), Prof. Xingjie Li(University of North Carolina at Charlotte), Ling Ye(China CDC in Daishan, Zhejiang), Long Chen(University of California, Irvine), Prof. Jiongming Yong(University of Central Florida) and Xinkang Chao(Mathworks). Wenbin Chen specially thank our colleague Prof. Rongmin Li who mentioned us to use the Gamma distribution. Wenbin Chen thanks Dr. Xu Chen’s group in the Winning Health Technology Group Company(winning.com.cn), who always believe and support us, he also thanks his wife Dr. Jie Xu, hers hometown is Wuhan.

We should thanks everyone who are fighting for Wuhan, specially we present our deep respects to Doctor Wenling Li and Prof. Nanshan Zhong.^1^.

## Footnotes

1 There may exists lots of typos and many references are note cited, since the paper is finished in a hurry and the first version is submitted at Feb. 17, 5:25am, 2020, after Xingjie Li helps me to modify the notes. Please let us know if you have any question and suggestions, please email us at.

